# Genome-wide by environment interaction study of stressful life events and hospital-treated depression in the iPSYCH2012 sample

**DOI:** 10.1101/2021.09.03.21262452

**Authors:** Nis P. Suppli, Klaus K. Andersen, Esben Agerbo, Veera M. Rajagopal, Vivek Appadurai, Jonathan R. I. Coleman, Gerome Breen, Jonas Bybjerg-Grauholm, Marie Bækvad-Hansen, Carsten B. Pedersen, Marianne G. Pedersen, Wesley K. Thompson, Trine Munk-Olsen, Michael E. Benros, Thomas D. Als, Jakob Grove, Thomas Werge, Anders D. Børglum, David M. Hougaard, Ole Mors, Merete Nordentoft, Preben B. Mortensen, Katherine L. Musliner

**Author notes:** **Corresponding author:** Katherine L. Musliner.

## Abstract

Researchers have long investigated a hypothesized interaction between genetic risk and stressful life events in the etiology of depression, but studies on the topic have yielded inconsistent results. We conducted a genome-wide environment interaction study in 18,532 depression cases from hospital-based settings and 20,184 population-representative non-cases from the iPSYCH2012 case-cohort study, a nationally representative sample identified from Danish national registers. Stressful life events including family disruption, serious medical illness, death of a first-degree relative, parental disability and child maltreatment were identified from population-based registers and operationalized as a time-varying count variable (0-4+). Hazard ratios for main and interaction effects were estimated using Cox regressions weighted to accommodate the case-cohort design. The analyses yielded three novel, genome-wide significant (P < 5 × 10^-8^) loci located in the ATP-binding cassette transporter C 1 (ABCC1) gene, in the A-kinase anchor protein 6 (AKAP6) gene, and near the Major facilitator superfamily domain 1 (MFSD1) gene, as well as 50 loci of suggestive significance. These top 3 hits did not replicate in a case-control sample of depression drawn from the UK Biobank. These results suggest that there may be individual genetic variants that confer risk for or protection against clinical depression only in the presence of stressful life events; however, replication in a sample with similar design and ascertainment methods is needed before any firm conclusions can be drawn. Future gene-by-stress research in depression should focus on establishing even larger collaborative genome-wide environment interaction studies to attain sufficient power.

## 1. Introduction

Major depression is a common, highly burdensome mental illness that effects as many as 21% of people at some point during their lifetimes ^1,2^. Studies suggest that major depression is around 30-40% heritable ^3^, meaning that a moderate amount of the population-level variability in major depression can be attributed to genetic factors. However, environment also plays an important role in determining who develops major depression and who does not. In particular, experiencing a stressful life event (SLE) in childhood or adulthood has been shown to increase depression risk ^4,5^. SLEs include childhood physical, sexual or emotional abuse, death of a relative, severe illness, divorce or separation, economic deprivation and forced exit from the workforce. Events can cause stress if they occur to the individual (e.g. child abuse, divorce, severe illness) or if they happen to a close relative, particularly during childhood (e.g. divorce or severe illness in a parent). Each of these events has been shown to be associated with increased risk for depression ^6,7^; however, the cumulative burden of stress is particularly relevant for determining depression risk. Studies have consistently shown that as the number of SLEs increases, risk for depression also increases, and individuals with over 4 SLEs experience depression risks 3-5 times that of individuals with no SLEs ^6,8,9^.

Historically there has been great interest in the possibility of an interaction between stressful life events and genetic liability as risk factors for depression. Such an interaction, if present, could lead not only to a better understanding of the underlying etiology of depression, but also potentially be useful for identifying individuals at particularly high risk for developing depression. An early twin study ^10^ found that risk for depression after an SLE was only elevated among individuals with high genetic liability. Subsequently, researchers selected candidate genes they believed were associated with depression risk and examined whether variants in these genes interacted with SLEs to predict depression ^11-19^. These studies yielded inconsistent results, with even meta-analyses reaching different conclusions regarding the validity of the associations ^20-27^. Research examining the interaction between polygenic risk scores and SLEs have also yielded inconsistent results, with some finding evidence for interaction ^28-30^ and some failing to do so ^29,31-33^.

The hypothesis-driven (i.e. candidate gene) approach for identifying specific variants associated with a given outcome has not been successful in psychiatric research ^34-36^. This led to the embrace of the genome-wide-association-study (GWAS) as a method for identifying variants associated with psychiatric disorders in a theoretically agnostic fashion. In a GWAS, single nucleotide polymorphisms (SNPs) in sufficient linkage disequilibrium to tag the entire genome are tested for association with the outcome of interest. Significance is evaluated based on an adjusted alpha level to avoid false positive results. This method has been highly successful in psychiatric genetics, and has led to the identification of over 100 variants associated with major depression at the genome-wide significant alpha level ^37^. Thus far, GWAS studies have failed to replicate any findings from candidate gene-by-stress interaction studies ^38^.

To our knowledge, four prior studies have used this theoretically agnostic, genome-wide approach to evaluate whether individual genetic variants interact with SLEs as risk factors for depressive symptoms measured using symptoms scales including the Beck Depression Inventory (BDI), the General Health Questionnaire (GHQ) and the Centers for Epidemiological Studies Depression Scale (CES-D): Dunn and colleagues ^39^ conducted a genome-wide environment interaction study (GWEIS) of depressive symptoms in a sample of 7,179 African American and 3,138 Hispanic/Latina women. They identified one genome-wide significant SNP in the African-American sample near the CEP350 gene (rs4652467, p = 4.10 × 10^-10^), however this association did not replicate. Ikeda and colleagues ^40^ conducted a GWEIS of depressive symptoms and SLEs in 1,088 individuals recruited from among employees of the Fujita Health University Hospital in Japan. The authors reported a significant interaction for a SNP near the BMP2 gene (rs10485715, p = 8.2 × 10^-9^), however no attempts were made to replicate this result. Otowa and colleagues ^41^ conducted a GWEIS of depressive symptoms and SLEs in 320 Japanese individuals, with no genome-wide significant results. Most recently, Arnau-Soler and colleagues conducted GWEISs of depressive symptoms and SLEs in 4,919 Europeans from the Generation Scotland cohort and 99,057 Europeans from the UK Biobank ^42^. The authors found 2 SNPs significant for interaction at the genome-wide level in the Generation Scotland sample: one near the PIWIL4 gene (p = 4.95 × 10^-9^) and one intronic to the ZCCHC2 gene (p = 1.46 × 10^-8^). They found no genome-wide significant hits in the UK Biobank, and the significant hits from the Generation Scotland Sample did not replicate in the UK biobank.

Most of these GWEIS studies had sample sizes that most likely left them underpowered to detect significant interaction results. In addition, the outcome of all of these studies was depressive symptoms rather than clinically defined major depression. Although depressive symptoms are highly genetically correlated with major depressive disorder ^43^, they nevertheless are a distinct outcome with, potentially, distinct associations with individual SNPs. Furthermore, all of these studies relied, out of necessity, on measures of SLEs that were retrospective and therefore potentially subject to recall bias ^44,45^. Finally, prior GWEIS studies were not able to account for the time-dependent nature of both SLEs and depression. SLEs can occur at multiple points during the lifespan, and analytic strategies that fail to account for this can potentially be subject to bias. GWAS has traditionally used logistic regressions to calculate odds ratios for the associations between individual SNPs and the odds of being a case. However, this approach does not measure risk for developing the disorder, which is arguably more useful from a clinical and public health standpoint ^46^. A different methodological approach is therefore needed to determine the associations between individual SNPs and risk for developing major depression, as well as potential interactions between SNPs and SLEs as risk factors for developing major depression.

Our aim in this study was to examine interactions between individual SNPs and a time-dependent, prospective measure of SLEs as risk factors for major depression in the general population. To accomplish this, we used data from the iPSYCH2012 case-cohort sample – a population-based cohort of individuals living in Denmark which includes information on psychiatric diagnoses from hospital-based settings. In addition, we also conducted a GWAS of depression using survival analysis rather than logistic regression as the underlying statistical methodology to examine the associations between individual SNPs and risk for developing depression in the general population.

## 2. Methods

### 2.1 Study design and sample

Data were drawn from the iPSYCH2012 study, which has a case-cohort design ^47^. In this design, the study sample is nested within a larger base population, and includes all cases from the full cohort but only a subset of non-cases. This reduces the cost and burden associated with collecting biological specimens (in the case of iPSYCH, DNA for genetic analysis). The subset used as the comparison group is typically a random sample of individuals drawn from the full cohort (i.e., the ‘subcohort). Because it is random, some cases will by chance be selected as part of the subcohort. The great benefit of this design over a nested case control design is that it enables the unbiased calculation of risk and hazard ratios, as in a cohort study ^48^. Because not all members of the full cohort are included, this design can be more efficient and cost effective than a cohort study particularly when the collection of biological samples is involved ^48-50^.

The iPSYCH2012 case-cohort sample includes a subcohort of 30,000 individuals selected randomly from the base population of all individuals born in Denmark between 1981-2005 who survived to their first birthday and had known mothers (i.e. the ‘subcohort’). To this random sample was added all additional individuals from the base population (N=56,189) who received a diagnosis of affective disorder, schizophrenia, autism, or ADHD between 1994-2012 in inpatient, outpatient or emergency room settings in Danish psychiatric hospitals (i.e. ‘cases outside the subcohort’). Records of psychiatric diagnoses are stored in the Danish Psychiatric Central Research Register (DCPRR) ^51^. Around 4% of individuals in the subcohort (n=1,188) also received one of the above psychiatric diagnoses, bringing the total number of psychiatric cases in the sample to 57,377. Biological material for DNA analysis was linked to information from national population-based registers using the unique, personal identification number assigned to all Danish citizens and legal residents since 1968 by the Danish Civil Registration System (DCRS) ^52^. The DCRS also includes parents’ personal identification numbers allowing establishment of all known first degree relatives (parents, siblings, half-siblings and offspring).

For this study, we selected all individuals in the iPSYCH2012 subcohort and all depression cases (ICD-10 codes F32-F33) outside the subcohort who were a) of European ancestry, b) successfully genotyped, and c) for whom follow-up data starting at age 10 years was available. We also removed at random 1 person from each pair of relatives (2^nd^ degree or closer, pi hat > 0.2). The final study sample included 38,716 individuals: 20,563 individuals from the subcohort (of whom 379 were also cases) and 18,153 depression cases outside the subcohort (total number of cases = 18,532).

### 2.2 Ethics statement

The iPSYCH2012 study was approved by the Danish Scientific Ethics Committee, the Danish Health Data Authority, the Danish Data Protection Agency, and Danish Newborn Screening Biobank Steering Committee. In accordance with Danish law, The Danish Scientific Ethics Committee has for this study waived the need for informed consent in biomedical research based on existing biobanks.

### 2.3 Measures

#### 2.31 Stressful life events

SLEs included death of a parent, sibling or child; serious medical illness in the participant or one of their first-degree relatives; family disruption due to divorce or separation, parental disability, and child maltreatment. SLE variables were obtained from Danish National population-based registers ^51,53,54^. A detailed description of how each SLE was measured is shown in Table 1. Dahl et al. ^6^ examined these events in the Danish registers and found that all were associated with depression risk individually, and that the number of SLEs was associated with depression in a dose-response fashion ^6^. Information on SLEs was combined into a time-varying count variable (0-4+ events), such that individuals contributed person time to the analyses within whichever category of SLE they were in at that time, and switched to contribute person time within a different SLE category when they experienced a subsequent SLE.

**Table 1.** Description of measures of stressful life events.

| **SLE variable** | **Description** |
| --- | --- |
| Death of a parent, sibling or child | Information on vital status (i.e. alive or dead) and date of death for all individuals residing legally in Denmark has been recorded in the Danish Civil Registration system (DCRS) since 1968. This register also includes information that can be used to link people with their first-degree relatives. We defined death of a parent, sibling or child as the date of death for that individual recorded in the DCRS. Death of a parent, sibling or child were handled as separate events. |
| Serious medical illness in the participant or their parent, sibling or child | Information on hospital treatment, including ICD diagnoses, for all individuals residing legally in Denmark is recorded in the Danish National Patient Register (DNPR). The DNPR includes all inpatient hospital visits since 1978 and all outpatient and emergency hospital visits since 1995. We defined serious medical illness based upon the Charlson Comorbidity Index (CCI). This index includes 19 severe somatic diseases all assigned a weighted score representing the associated 1-year relative risk of dying. Diagnoses in the CCI have a very high accuracy in the DNPR. Individuals were coded as 0, 1, or 2+ based on their CCI score. For the participant and first-degree relatives, we considered dates of changes in CCI score from 0 to 1, and 1 to 2+ as separate events. |
| Family disruption | The annual cohabitation status (defined on January 1 each year) for all individuals residing legally in Denmark has been recorded in the DCRS since 1980. We defined family disruption in childhood as all instances where a person was registered as a child in a family with two cohabitating adults – of whom at least one was the legal parent of the child – who ceased to live together. Family disruption due to death of a parent was not included in this variable. Family disruption in adulthood was defined as the participant themselves ceasing to cohabit with an adult to whom they were married or, as many partnered adults in Denmark choose not to marry, with whom they had a child. Date of family disruption was defined as January 1 of the year in which cohabitation status changed. |
| Parental disability | Employment status for all individuals residing legally in Denmark has been registered each year on November 1 in the Danish register on personal labour market affiliation since 1981. Denmark offers a disability pension to individuals who are unable to work due to physical or mental disability. We measured date of parental disability as November 1 in the first year that a participant's mother or father was registered as receiving a disability pension. Disability in the mother and father were treated as separate events. |
| Child maltreatment | Information on child maltreatment was obtained from the DNPR. We defined child maltreatment as the date at which the participant received one of the following ICD-10 codes: T74.0 - neglect or abandonment; T74.2, Z61.4, Z61.5 – sexual abuse; T74.1 – physical abuse; T74.3 – psychological abuse; T74.8, T74.9 – other or unspecified maltreatment syndromes. |

#### 2.32 Genetic data

DNA was obtained from blood spots collected at birth as part of routine clinical screening and stored in the Danish Newborn Screening Biobank ^55^. Bloodspots were located for 80,422 members (93%) of the iPSYCH2012 sample. Samples were genotyped at The Broad Institute of Harvard and MIT (Cambridge, MA, USA) in 23 waves using the Infinium PsychChip v1.0 array (Illumina, San Diego, CA, USA). Quality control (QC) and imputation were performed using the RICOPILI pipeline ^56^. The filtering process excluded variants with call frequency < 0.98 or a Hardy-Weinberg Equilibrium *P* value < 1 × 10^-6^. Ninety percent (N=77, 639) of the sample passed QC.

### 2.4 Analyses

Main and interaction effects for the associations between individual SNPs, SLEs and depression were estimated using a series of Cox regressions weighted according to the Prentice method to accomodate the case-cohort design ^48,49^. Persons in the study sample were followed from age 10 until depression diagnosis, death, emigration, or December 31, 2012, whichever came first. The underlying time-metric was age in days. The time-dependent SLE count variable was analyzed as a continuous variable. All analyses were adjusted for sex, birth year and the first 5 ancestral principal components. Wald statistics were used to test for interaction. Analysis were conducted in R (version 3.1.2; R Foundation, Vienna, Austria). Regional visualizations of results from GWEIS analyses were plotted with LocusZoom ^57^.

There are approximately 11 million directly genotyped and imputed SNPs available for members of the iPSYCH2012 sample. However, according to Danish law, some register-based data is available only at dedicated servers at Statistics Denmark (Copenhagen, DK). Because this study includes variables that can only be accessed through these servers, we were required to conduct the analyses in a Windows environment, which created some computational challenges that made it impossible to run our GWAS and GWEIS analysis in the full set of 11 million SNPs. To get around these challenges, we conducted our GWEIS of SLEs and depression in two stages: first, we selected a subset of SNPs where MAF > .01 and missing rate < .1. From there, we conducted linkage disequilibrium (LD) pruning with various r^2^ thresholds and found that an *r*^*2*^ value of 0.7 left us with 496,162 high quality SNPs distributed across the genome. These SNPs were then uploaded onto the Statistics Denmark servers and merged with the register-based data for GWAS and GWEIS analysis. Based on the GWEIS analysis using these 496,162 SNPs, we identified all SNPs with interaction p-values below the commonly used genome-wide suggestive threshold (*P* < 1 × 10^-5^). We then went back to the original sample of 11 million SNPs and identified all those SNPs located 500 kb upstream or downstream of the suggestive SNPs and uploaded them onto the server at Statistics Denmark. This enabled a second stage of analysis in which there was dense coverage of the areas with suggestive evidence for interaction. For this second stage, statistical significance was evaluated at the genome-wide significant alpha level of *P* < 5 × 10^-8^. Given the actual number of SNPs included in our GWEIS and the fact that the second stage of SNP selection specifically aimed to increase coverage of specific genomic areas, we posit P < 5 × 10^-8^ to be a conservative threshold.

#### 2.41 Replication attempt

We attempted to replicate our top findings in a case-control sample of depression drawn from the UK Biobank ^58^. The UK Biobank includes more than 500,000 persons aged 40–69 at recruitment and holds a variety of biological measurements, lifestyle indicators and biomarkers, including genome-wide genotype data on all participants. The current replication analyses were based on a sample of 73,258 genetically unrelated persons of European ancestry (22,880 depression cases and 50,378 controls) ^59^ for whom SNP data as well as information on trauma exposure were available ^60^. Lifetime depression was assessed with questions from the Composite International Diagnostic Interview. Trauma exposure was operationalized as a dichotomous variable based on self-report of severe trauma experiences in childhood and adulthood. We tested for interaction between the dichotomous trauma exposure and all available SNPs located within +/- 500 kb of the most significant SNP from each of the three genome-wide significant loci identified in the iPSYCH2012 GWEIS. In total, 7745 SNPs were tested for interaction using PLINK2a ^61^. We assessed the number of independent loci tested for interaction at varying r^2^ (0.1-0.5) and differently sized windows (250-3000Kb) yielding 443 to 1252 independent loci (See Table 2).

**Table 2.** Estimated number of independent loci tested for interaction in the replication analyses.

| Independent loci | Kb | r<sup>2</sup> | Bonferroni adjusted p values |
| --- | --- | --- | --- |
| 1252 | 250 | 0.5 | 3.99 x 10<sup>-5</sup> |
| 1232 | 3000 | 0.5 | 4.06 x 10<sup>-5</sup> |
| 1073 | 250 | 0.4 | 4.66 x 10<sup>-5</sup> |
| 1051 | 3000 | 0.4 | 4.76 x 10<sup>-5</sup> |
| 866 | 250 | 0.3 | 5.77 x 10<sup>-5</sup> |
| 842 | 3000 | 0.3 | 5.94 x 10<sup>-5</sup> |
| 687 | 250 | 0.2 | 7.28 x 10<sup>-5</sup> |
| 653 | 3000 | 0.2 | 7.66 x 10<sup>-5</sup> |
| 493 | 250 | 0.1 | 1.01 x 10<sup>-4</sup> |
| 443 | 3000 | 0.1 | 1.13 x 10<sup>-4</sup> |

## 3. Results

### 3.1 Sample characteristics

Sample characteristics are shown in Table 3. Collectively, participants in the sample contributed a total of 395,331 person-years of follow-up, with an average follow-up time of 10 years (standard deviation = 5 years). Depression cases inside and outside the population-based random subcohort showed similar characteristics. Sixty-none percent of cases and 49% of subcohort members were female. Mean age at first depression diagnosis in cases was 19.6-19.7 years (*SD* = 4.1-4.2 years). SLEs were common – by age 10, 48-49% of the cases and 39% of subcohort members had experienced at least one SLE.

**Table 3.** Sample characteristics.

| Characteristic | MD cases outside the subcohort<br>(N = 18,153) | MD cases inside the subcohort<br>(N = 379) | Non-cases from the subcohort<br>(N = 20,184) |
| --- | --- | --- | --- |
|  | N (%) | N (%) | N (%) |
| Gender |  |  |  |
| Female | 12,430 (68.5) | 263 (69.4) | 9,848 (48.8) |
| Male | 5723 (31.5) | 116 (30.6) | 10,336 (51.2) |
| Birth cohort |  |  |  |
| 1981-1985 | 5953 (32.8) | 126 (33.3) | 3 585 (17.8) |
| 1986-1990 | 6670 (36.7) | 150 (39.6) | 4 570 (22.6) |
| 1991-2002 | 5530 (30.5) | 103 (27.2) | 12 029 (59.6) |
| >1 SLE before age 10 years | 8,712 (48.0) | 185 (48.8) | 7,857 (38.9) |
| Age at first MD diagnosis, years<br>(mean, SD) | 19.7 (4.2) | 19.6 (4.1) | NA |

### 3.2 GWAS results

Figure 1 shows results from GWASs examining the main effects of 496,162 SNPs on hazard of depression (Figure 1a) and hazard of experiencing at least one SLE (Figure 1b). The GWAS of the risk for developing depression yielded one genome-wide significant hit (rs7700661, *P* = 1.99 × 10^-8^) and 52 suggestive hits (*P* < 1 × 10^-5^) (Figure 1a). No individual SNPs reached genome-wide or suggestive significance for the main effect of SNPs on hazard of SLEs (Figure 1b).

**Figure 1.**
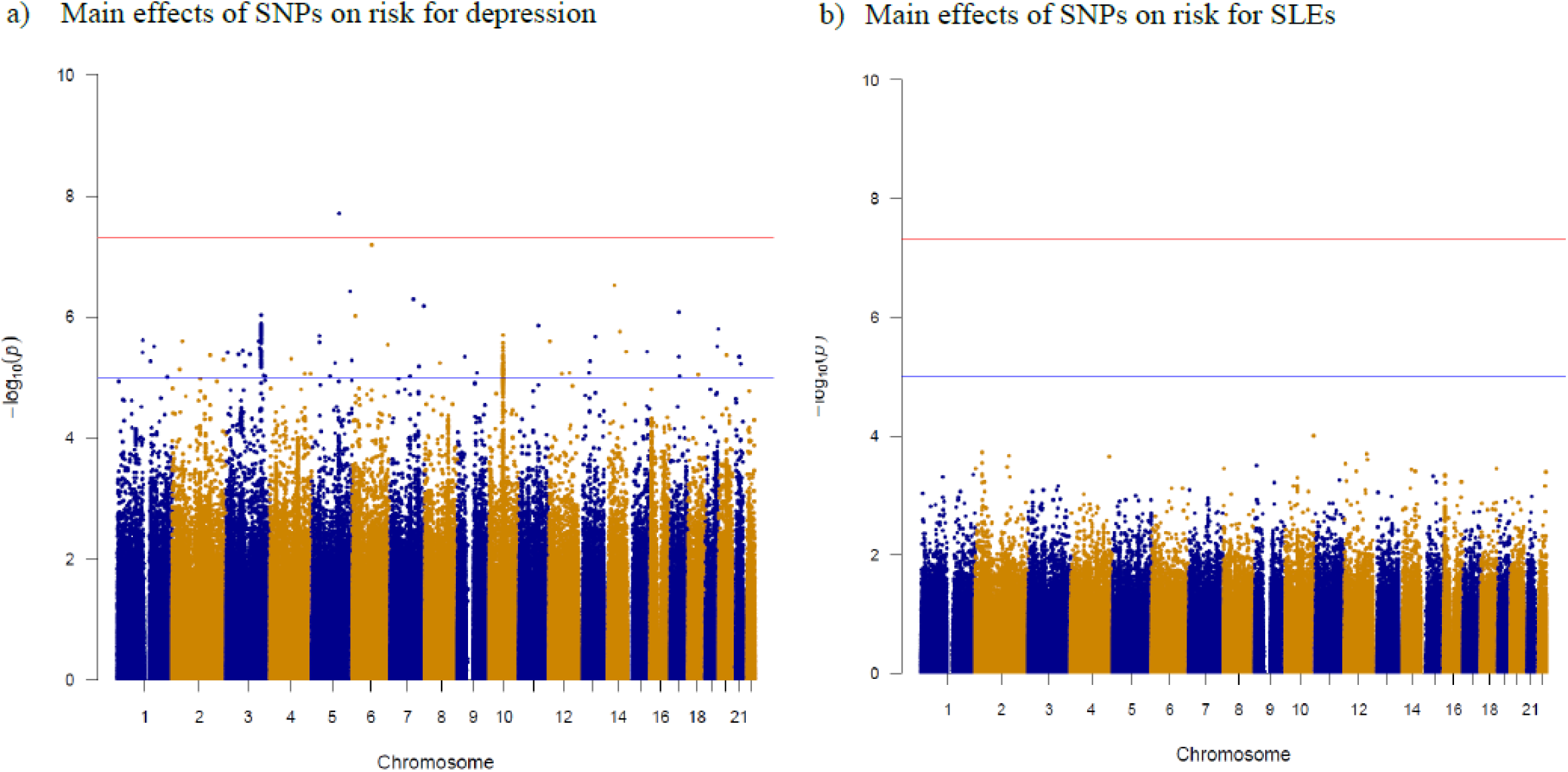
Manhattan plots for main effects of 496,162 SNPs on risk for depression and stressful life events in 18,532 depression cases and 20,563 random subcohort members. Note. Panel A shows main effects of 496,162 individual SNPs on risk for depression diagnosis in hospital-based settings in Denmark from 1995-2012. Panel B shows main effects of 496,162 individual SNPs on risk for experiencing at least one stressful life event.

### 3.3 GWEIS results

The GWEIS analysis of 496,162 SNPs yielded 60 SNPs that reached suggestive significance (*P* < 1 × 10^-5^, Table 4). After re-running the GWEIS including all SNPs located within 500 Kbs of these 60 SNPs, three independent loci reached genome-wide significance (Figure 2). Hazard ratios for the 3 top hits are shown in Figure 3, and region plots in Figure 4. The top hit, rs56076205 (*P* = 3.7 × 10^-10^), was located in an intron of the ATP-binding cassette transporter C 1 (ABCC1) gene. Compared with homozygotes for the major allele and heterozygotes, homozygotes for the minor allele (minor allele frequency (MAF)= 0.07) showed a pattern of increasing hazard ratio (HR) for depression with increase number of SLEs (see Figure 3a). The second hit, rs3784187 (*P* = 1.2 × 10^-8^), was located in an intron of the A-kinase anchor protein 6 (AKAP6) gene. For this SNP, homozygotes for the minor allele (MAF 0.06) showed a negative interaction such that as SLEs increased, risk for depression decreased (see Figure 3b). The final hit, rs340315 (*P* = 4.5 × 10^-8^), was located near the Major facilitator superfamily domain 1 (MFSD1) gene. Homozygotes for the minor allele (MAF 0.31) showed a similar pattern to the first hit, in which HR for depression increased as number of SLEs increased (see Figure 3c).

**Table 4:**
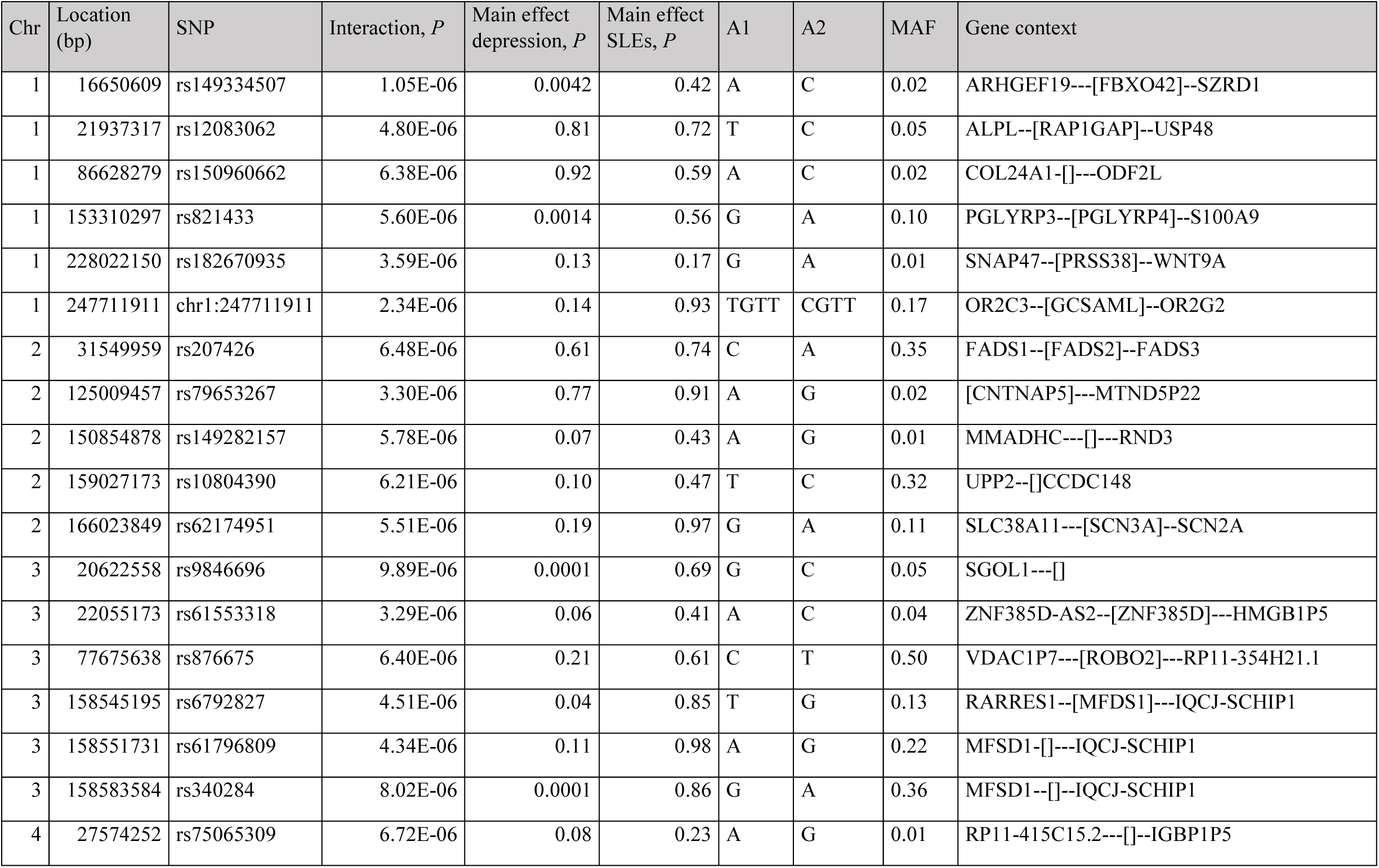
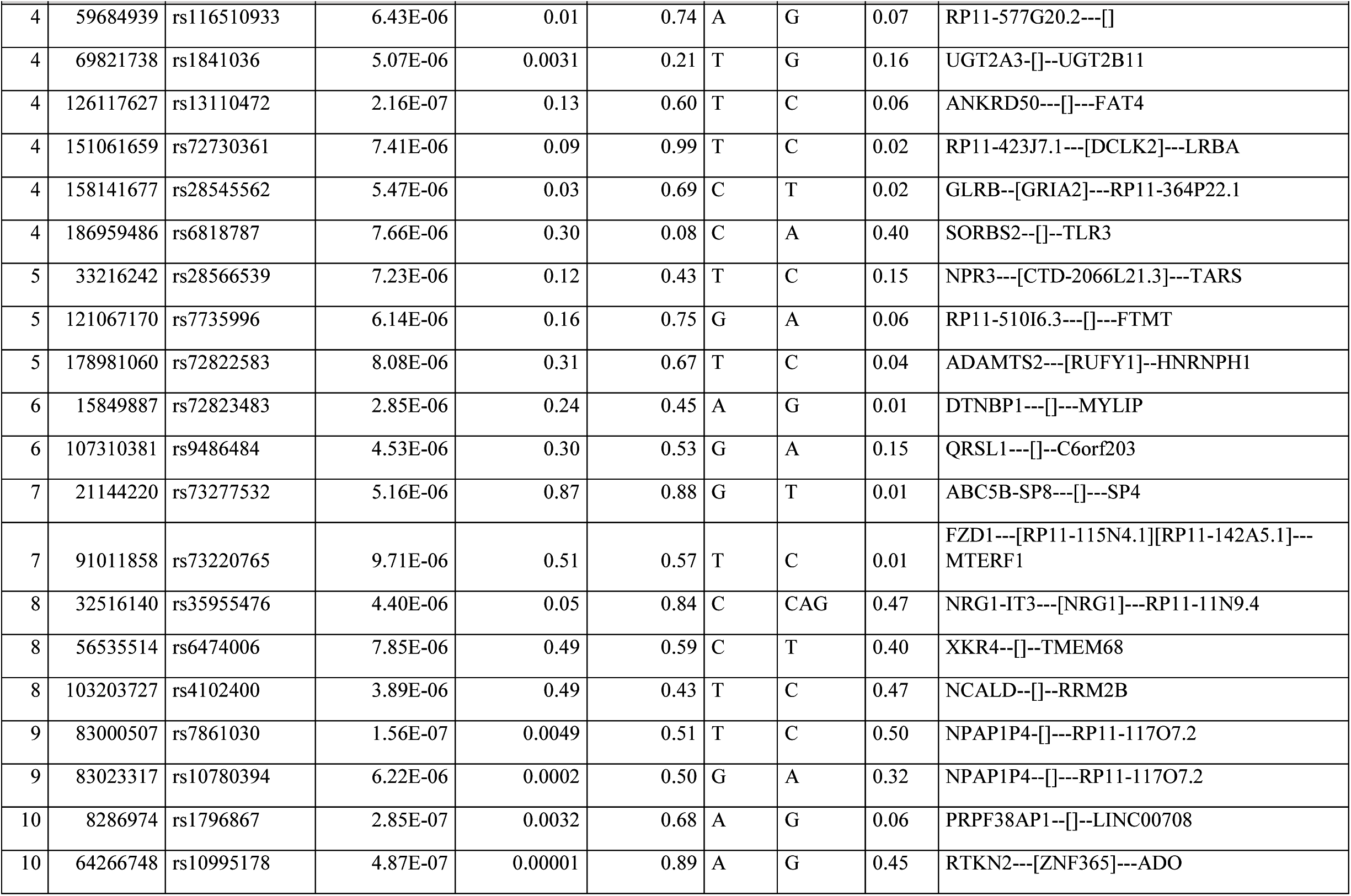
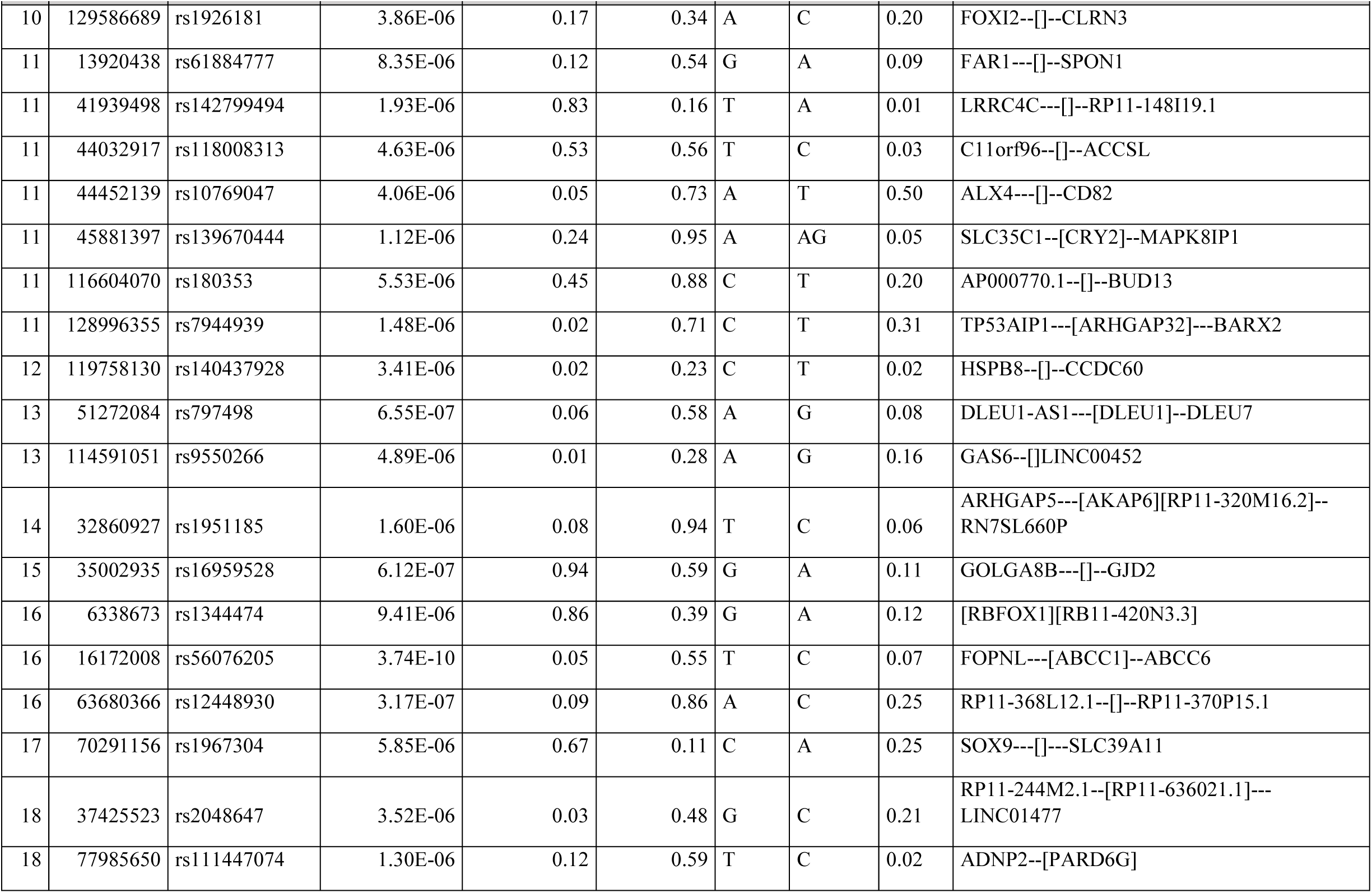
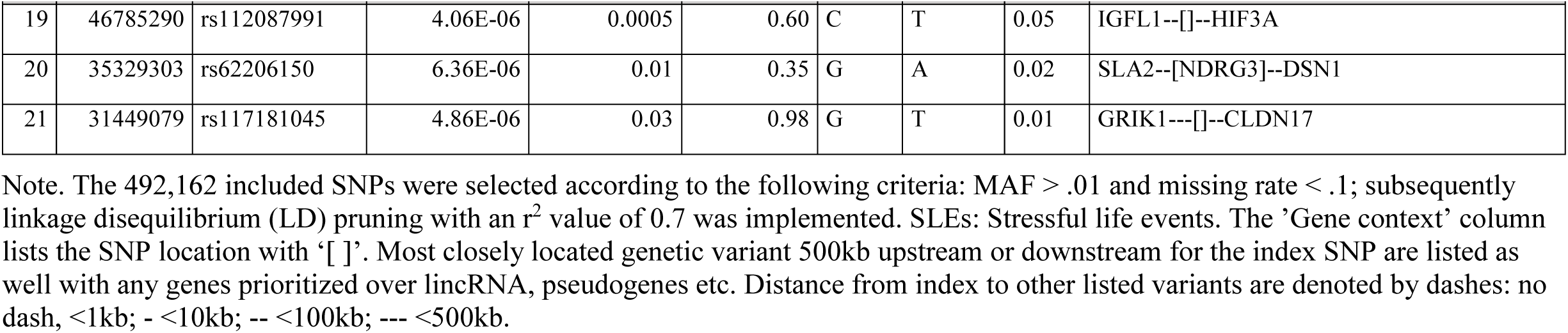
60 SNPs passing genome-wide suggestive threshold (*P*<1×10^-5^) for interaction with stressful life events on risk for depression tested among 496,162 SNPs in 18,532 depression cases and 20,563 random subcohort members.

**Figure 2.**
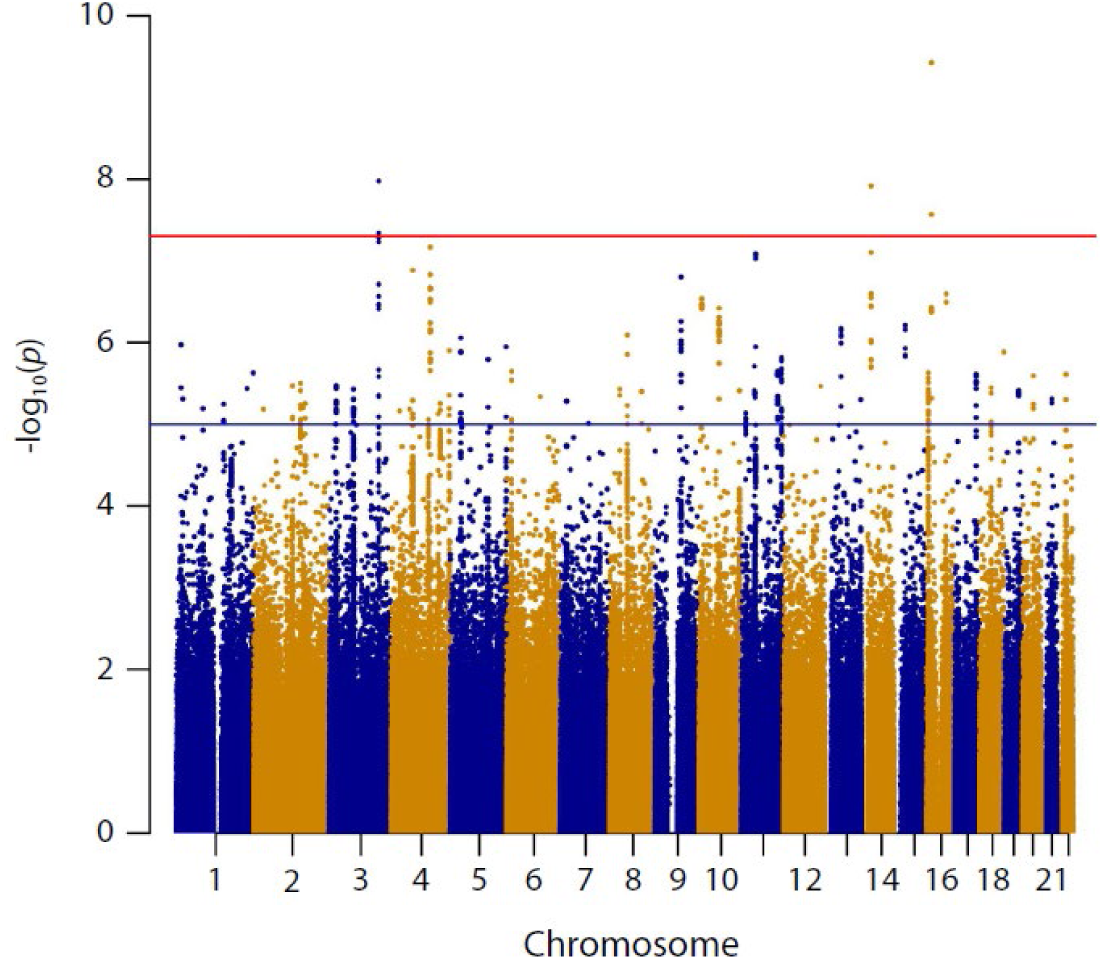
Manhattan plot of genome-wide environment interaction analyses based on 18,532 depression cases and 20,563 random subcohort members. Note. The figure presents results of a genome-wide environment interaction study (GWEIS) conducted in two stages: In stage 1, a GWEIS was conducted using 496,162 SNPs distributed across the genome. In stage 2, all SNPs located 500kb up- or downstream from 60 SNPs with p values < 10^-5^ in stage 1 were added to the analyses. The Manhattan plot shows results from both stages.

**Figure 3.**
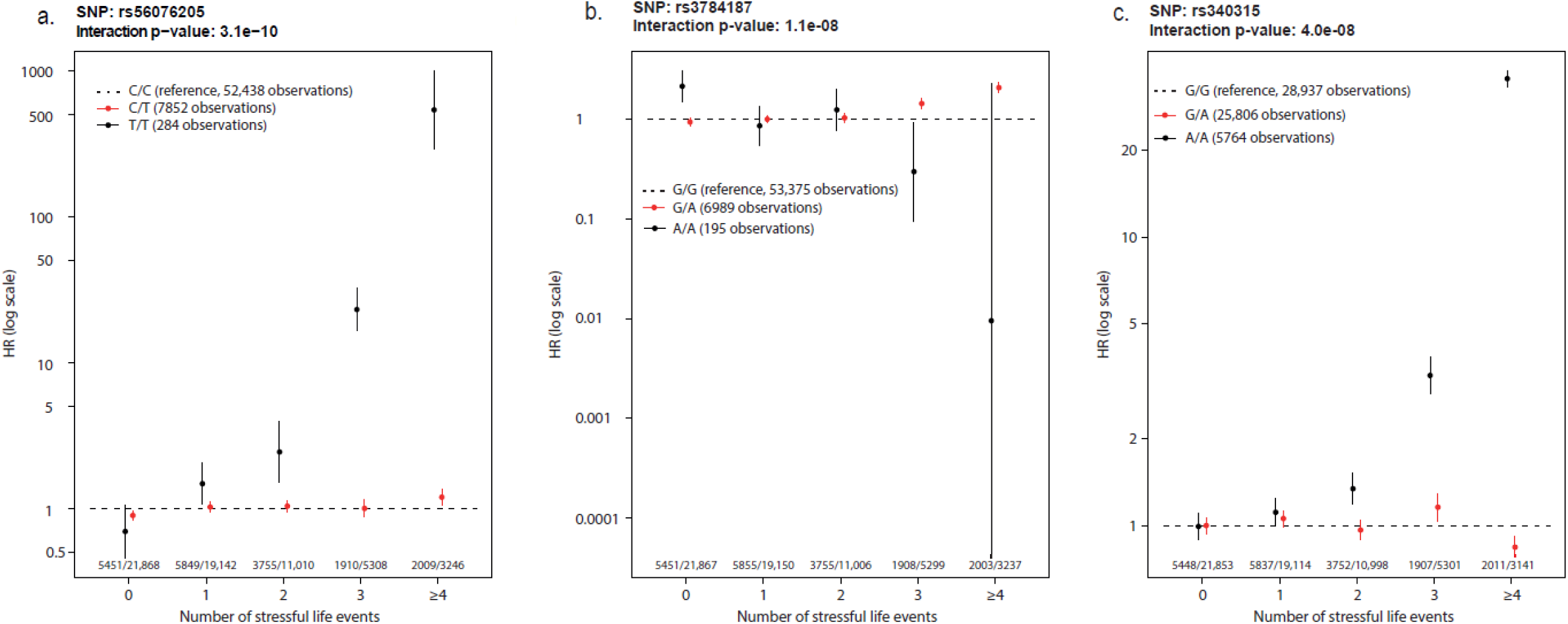
Interaction effects for stressful life events and top SNPs from 3 genome-wide significant loci. Note. For each SNP, the hazard ratio (HR) for depression is plotted by number of stressful life events. Vertical bars represent 95% confidence intervals. Hazards were compared within each level of stress life events with major allele homozygotes as reference. Wald statistics were used to test interactions, comparing linear trends for HR between genotypes. The small differences in the total number of observations are due to differences in the number of persons successfully genotyped for each SNP. Due to the time-varying nature of the stressful life events variable, study participants could contribute person-time for different numbers of stressful life events. Therefore, the total number of observations exceeds the total number of participants in the study.

**Figure 4.**
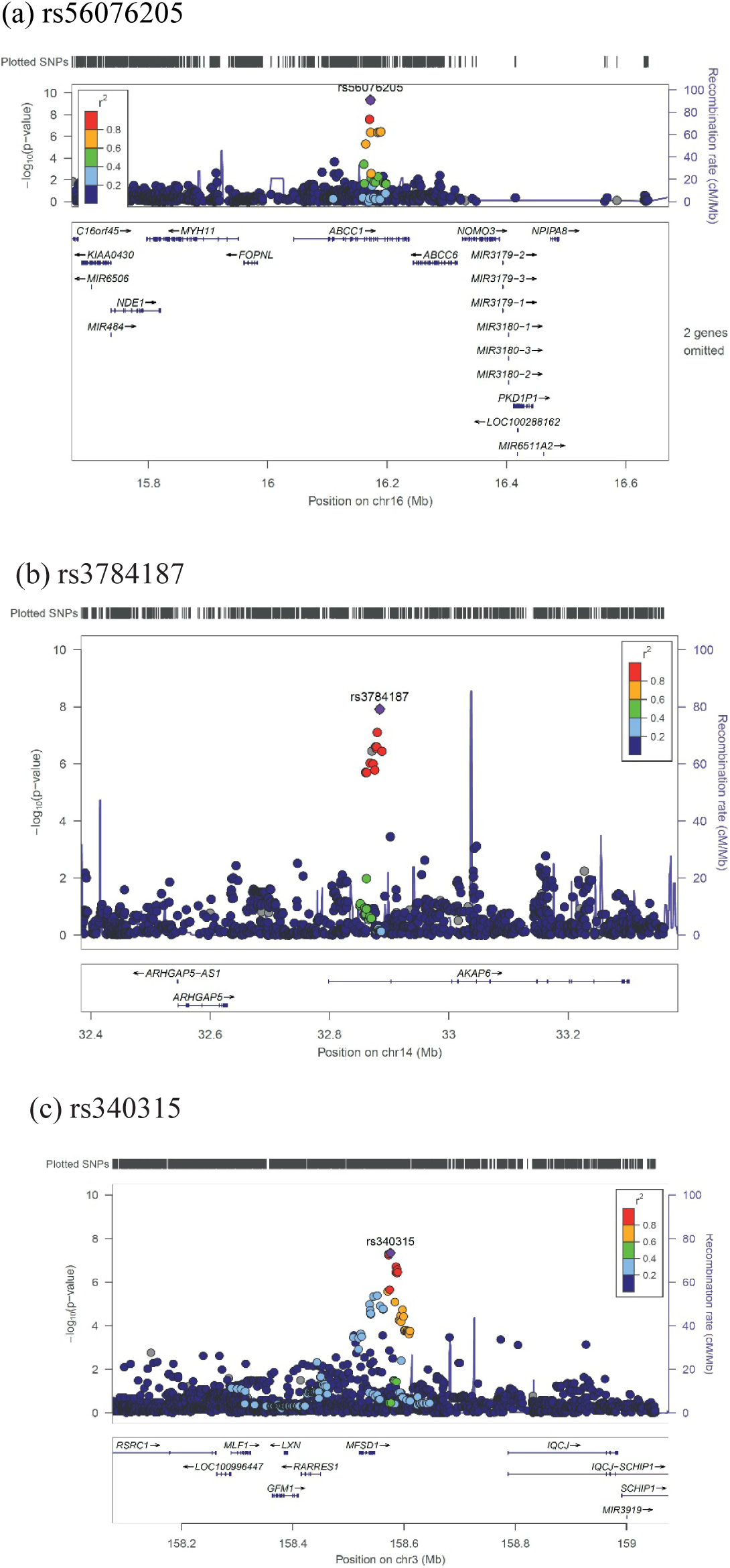
Region plots for 3 top hits from a genome-wide by environment interaction study based on 18,532 depression cases and 20,563 random subcohort members. Note. Color of dots indicate the linkage disequilibrium (r^2^) of SNPs with the top SNP of each loci. The r^2^ was based on the 1000 Genomes November 2014 EUR population.

### 3.4 Analysis of top SNPs in UK Biobank

None of the three top SNPs were statistically significant in the replication attempt using UK Biobank data (rs56076205, *P* = 0.87; rs3784187, *P* = 0.93; rs340315, *P* = 0.71). The most significant interactions involved the following SNPs: rs190869692 (*P* = 3.2 × 10^-5^) in the *ABCC1* gene 38,653 bases upstream from the iPSYCH2012 hit in the same gene (r^2^ = 0.002; *P* = 0.58); rs111284027 (*P* = 9.4 × 10^-5^) in the *ARHGAP5* gene 259,273 bases downstream from our hit in the *AKAP6* gene (r^2^ = 0.003; *P* = 0.44); rs146472082 (*P* = 5.1 × 10^-5^) in the *RARRES1* gene 155,569 base pairs downstream from our hit near the *MFSD1* gene (r^2^ = 0.053; *P* = 0.0011) (See Figure 5). Thus, all three SNPs identified in the replication analyses represented independent loci from the three genome-wide significant loci identified in the iPSYCH2012 GWEIS.

**Figure 5.**
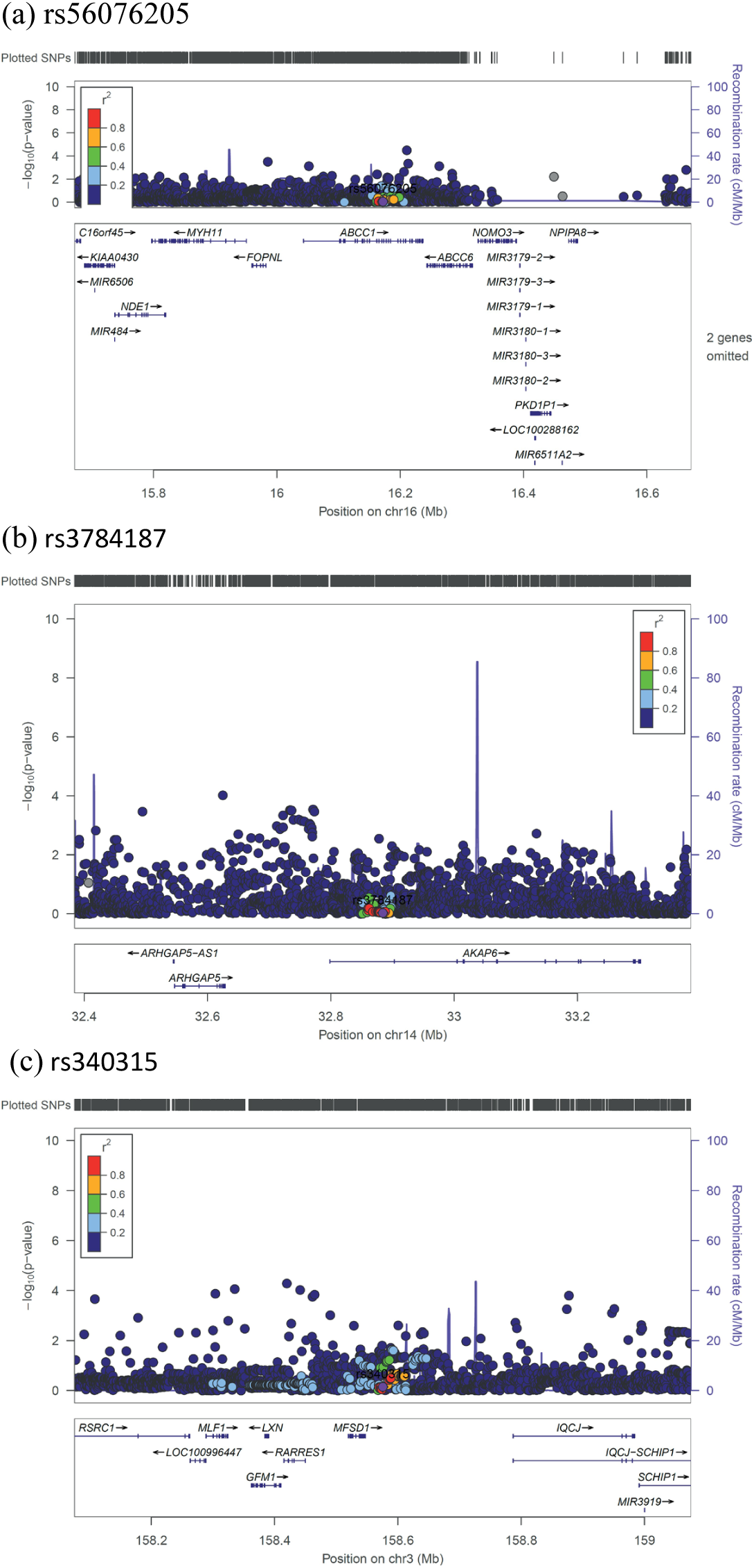
Region plots for the three most significant SNPs in the UK Biobank replication attempt. Note. SNPs identified with genome-wide significant interactions in analyses of iPSYCH data were used as index SNPs in the above regional plots of the UK Biobank replication attempt. Color of dots indicate the linkage disequilibrium (r^2^) of SNPs wiht the top SNP at each loci identified in the interaction analysis based on iPSYCH2012. The r^2^ was based on the 1000 Genomes Nov 2014 EUR sample

## 4. Discussion

In this study, we report results from the first comprehensive, population-based genome-wide environment interaction study investigating the interaction between individual SNPs and a time-varying measure of stressful life events (SLEs) as risk factors for a diagnosis of depression treated in inpatient, outpatient or emergency room settings. The GWEIS yielded genome-wide significant effects in three independent loci located in the *ABCC1, AKAP6* and *MSFD1* genes, as well as 50 hits of suggestive significance. The *ABCC1* is known as a multidrug resistance protein and has a range of commonly used drugs as substrate ^62^. Mice studies report a strong influence of *ABCC1* on cerebral accumulation of beta-amyloid ^63^. One study also reported that St. John’s wort (*Hypericum perforatum)*, which has an antidepressant effect ^64^, increased the transport efficacy of *ABCC1* ^*65*^. This interaction is compatible with the idea of ‘plasticity’ genetic variants associated with stronger responses to both positive—in this study the absence of SLEs—and negative environmental exposures ^66^. Theoretically, it would be difficult to identify a main effect of genetic variants showing such plasticity. The protein transcribed from the *AKAP6* gene is involved in intracellular signaling in the protein kinase A pathway and *AKAP6* is highly expressed in brain tissue, cardiac and skeletal muscle ^67^. In 2015, a meta-analysis from the CHARGE consortium found a genome-wide significant association between a SNP in the *AKAP6* gene and general cognitive functioning ^68^. The *MFSD1* is a membrane-bound solute carrier present in a wide range of human tissues ^67^. A recent mice study reported the *MFSD1* to be abundant in the plasma membrane of neurons ^69^. Further, the study found alterations in gene expression in response to environmental stress in the form of starvation.

None of the top hits from the iPSYCH2012 GWEIS analysis were significant in the UK Biobank. Although a successful replication of one or more of these hits would have provided convincing support for their validity, the absence of successful replication in this case is difficult to interpret due to the large differences between iPSYCH2012 and UK Biobank in terms of sampling, measurement, and design. The fact that different statistical methods were used (survival analysis vs. logistic regression) could also be a contributing factor. However, it is not straightforward to isolate the impact of the statistical method alone, because conducting a logistic regression in our own sample would require us to make substantial changes to the design and sample composition. Thus it would be difficult to tell if any difference in the results were due to the different statistical method or the different design. Ultimately, a more solid conclusion regarding the validity of these hits will only be obtained through a replication attempt in a sample with comparable measurement and design. Such a sample will hopefully be available in the near future, but until that time, these hits should be considered suggestive.

To our knowledge, this is the largest single-sample GWEIS conducted to date examining the interaction between individuals variants and SLEs. Nevertheless, the presented analyses are still likely underpowered to detect most single SNP gene-environment interactions ^70^. For years, GWASs were similarly underpowered to detect significant SNPs, until the development of large-scale international consortia allowed for the accumulation of enough samples to pass the inflection point for consistent findings ^71^. In comparison, the study of gene-environment interaction in psychiatric disorders has only begun to enter into it’s ‘big data’ phase. The requirement for assessment of a complex, composite environment exposure in in the large study populations inherently required to study interactions makes these studies challenging endeavors. Extrapolating from the history of GWASs in psychiatry, we believe that the inflection point for studies of gene-environment interaction will only be reached through international collaborations that combine studies with information on genetic variation and environment exposures.

### 4.1 Methodological considerations

The following are additional methodological aspects of the study that should kept in mind when interpreting these results. First, the oldest depression cases in the iPSYCH2012 sample were diagnosed by age 30. As such, they represent a cohort of early onset depression cases and therefore these results may not generalize to individuals who develop depression at older ages. Second, the depression cases in iPSYCH are all identified in hospital-based settings, therefore these results may not generalize to individuals with untreated depression or individuals treated solely by their primary care doctors, who make up the majority of depression cases in Denmark ^72^. Fourth, although some of the SLEs included in this study are measured with high accuracy (e.g. death of a relative), others, particularly child maltreatment, are measured less accurately because they are based solely on register data. It is sadly very likely that some individuals in the sample experienced child maltreatment that was never recorded in the register, although the opposite (that individuals registered as having experienced child maltreatment did not experience it) is unlikely to be true. Third, we included a diverse range of stressful events in our study. Consequently, it is possible that some observed interactions relate to very specific types of stressful life events. For example, it is plausible that risk for depression in relation to somatic disease is associated with the seriousness of the course of disease. Therefore, genetic variants associated with prognosis and/or treatment response could emerge as part of gene-environment interaction in the present study. E.g. the ABCC1 has a range of anti-cancer and anti-HIV drugs as substrates, thus rendering somatic treatment less effective thereby possibly increasing risk for depression.

### 4.2 Conclusion

In this population-based cohort of European ancestry, we identified 3 novel genetic loci that interacted with a time-varying measure of stressful life events to predict hospital-treated depression at a genome-wide significant level, and over 50 additional loci with suggestive evidence for interaction. These results await confirmation via replication in a study with sufficiently similar measures and design. Future gene-by-stress research in depression should focus on efforts to establish large collaborative genome-wide environment interaction studies to generate sufficient statistical power to identify significant variants.

## Supporting information

STROBE checklist

## Data Availability

Data referred to in the manuscript are available from Statistics Denmark. Restrictions apply to the availability of these data, which were used under license for this study. For more information on accessing the data see www.dst.dk.

## Acknowledgments and Disclosures

The iPSYCH project is funded by the Lundbeck Foundation (grant numbers R102-A9118, R155-2014-1724 and R248-2017-2003) and the universities and university hospitals of Aarhus and Copenhagen. Katherine Musliner is funded by a postodoc fellowship from The Lundbeck Foundation (R303-2018-3551). Genotyping of the iPSYCH2012 samples was supported by grants from the Lundbeck Foundation, the Stanley Foundation, the Simons Foundation (SFARI 311789), and the National Institutes of Mental Health (NIMH 5U01MH094432-02). The Danish National Biobank resource is supported by the Novo Nordisk Foundation. The authors gratefully acknowledge The Broad Institute for genotyping. Initial genetic analyses were performed on the GenomeDK HPC facility suppored by Center for Genomics Personalized Medicine and iSEQ, Aarhus University. The UK Biobank (Project ID 16577) represents independent research supported by the National Institute for Health Research (NIHR) Biomedical Research Centre at South London and Maudsley NHS Foundation Trust and King’s College London. The views expressed are those of the authors and not necessarily those of the NHS, the NIHR or the Department of Health and Social Care. High performance computing facilities at the NIHR BRC were funded with capital equipment grants from the GSTT Charity (TR130505) and Maudsley Charity (980).

Thomas Werge has served as scientific advisor to H. Lundbeck A/S. Gerome Breen has received consultancy and speaker fees from Eli Lilly, Otsuka, and Illumina. The rest of the authors have no conflicts of interest to report.

